# Time-Dependent ECG-AI Prediction of Fatal Coronary Heart Disease

**DOI:** 10.1101/2023.10.11.23296910

**Authors:** L. Butler, A. Ivanov, T. Celik, I. Karabayir, L. Chinthala, S. M. Tootooni, B. C Jaeger, A. Doerr, D. D. McManus, L. R. Davis, D. Herrington, O. Akbilgic

## Abstract

**Background:** Sudden cardiac death (SCD) affects >4 million people globally, and ∽300,000 yearly in the US. Fatal coronary heart disease (FCHD) is used as a proxy to SCD when coronary disease is present and no other causes of death can be identified. Electrocardiographic (ECG) artificial intelligence (AI) models (ECG-AI) show promise in predicting adverse coronary events yet their application to FCHD is limited.

**Objectives:** This research aimed to develop accurate ECG-AI models to predict risk for FCHD within the general population using waveform 12- and single-lead ECG data as well as assess time-dependent risk.

**Methods:** Standard 10-second 12-lead ECGs sampled at 250Hz, demographic and clinical data from University of Tennessee Health Science Center (UTHSC) were used to develop and validate models. Eight models were developed and tested: two classification models with convolutional neural networks (CNN) using 12- and single-lead ECGs as inputs (12-ECG-AI and 1-ECG-AI, respectively) and six time-dependent cox proportional hazard regression (CPHR) models using demographics, clinical data and ECG-AI outputs. The dataset was split into 80% for model derivation, with five-fold cross-validation, and 20% holdout test set. Models were evaluated using the AUC and C-Index. Correlation of predicted risks from the 12-lead (12-ECG-AI) and single-lead (1-ECG-AI) CNN models was assessed.

**Results:** A total of 50,132 patients were included in this study (29,093 controls and 21,039 cases) with a total of 167,662 ECGs with mean age of 62.50±14.80years, 53.4% males and 48.5% African-Americans. The 12- and 1-ECG-AI models resulted AUCs=0.77 and 0.76, respectively on the holdout data. The best performing model was C12-ECG-AI-Cox (demographics+clinical+ECG) with no time restriction AUC=0.85(0.84-0.86) and C-Index= 0.78(0.77-0.79). 2-year FCHD risk prediction reached AUC=0.91(0.90-0.92). The 12-/1-ECG-AI models’ predictions were highly correlated (R^2^ = 0.72).

**Conclusion:** 2-year risk for FCHD can be predicted with moderate accuracy from ECG data alone. When combined with other data, a very high accuracy was obtained. High correlation between single-lead and 12-lead ECG models infer opportunities for screening larger patient populations for FCHD risk.

**Graphical Abstract:** 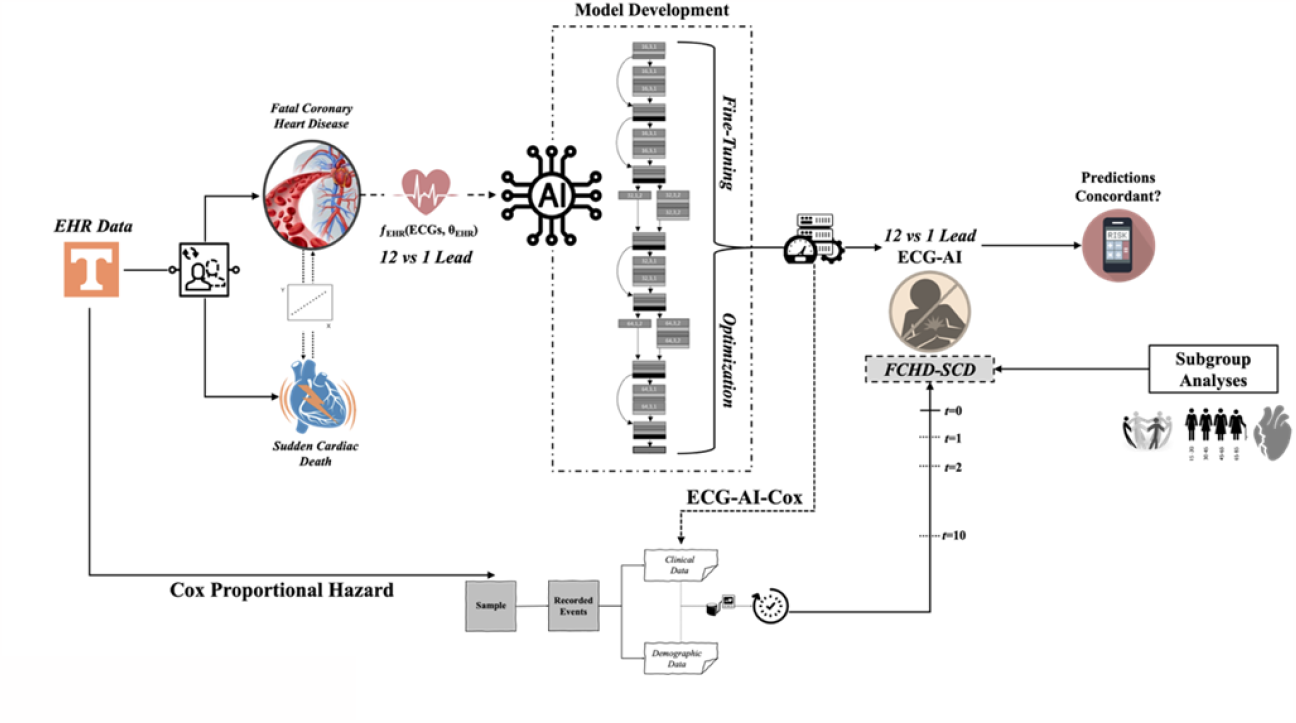

## 1. Introduction

SCD is an unexpected and sudden cardiovascular collapse with ∽80% of cases resulting in death before reaching the hospital^1,2^. SCD is amongst the most common causes of death, affecting more than 4 million people globally, with an estimated 300,000 events yearly in the United States alone^3^ with increasing prevalence in adults aged 30-40 years^3,4^.

Recent literature suggest that fatal coronary heart disease (FCHD) is a proxy to SCD^5,6^. There have been developments in potential primary prevention when risk of FCHD or SCD have been detected, often relying on prevention or treatment of ischemic events, heart failure (HF), coronary artery disease (CAD), hypertrophic cardiomyopathy (HCM) or arrhythmias (including fatal arrhythmias). In most cases, the attributable prevention strategy is the use of implantable cardioverter defibrillators however there are cases where the underlying cardiovascular irregularity is clinically addressed via appropriate pharmacological therapies such betablockers for long QT syndrome^7^ and non-ischemic evaluation for those with anomalous coronary arteries^8,9^. However, early detection and prediction of the event still remains challenging since risk modification strategy requires warning ^10,11^ to help reduce chance of SCD event using an appropriate intervention strategy.

Artificial intelligence (AI) methods have become assets in detecting and predicting risk for different cardiovascular diseases due to the large availability of clinical data^12,13^. There is potential for the development of AI models for pre-screening during any clinical service to assess the risk of FCHD/SCD at an early enough stage to allow for prevention or timely therapeutics. Current research has tried to predict FCHD risk using clinical risk factors^14^, ECG features as biomarkers^5,15^, cardiac imaging^16,17^ and have also made available a 5-year SCD risk calculator which is specific to people with hypertrophic cardiomyopathy^14^ but these have limitations attributed to prediction performance, small sample sizes or require additional resources, which might not be accessible to everyone.

Prior studies have shown that ECG features (e.g., QRS amplitude, QRS duration, etc.) relate to risk for SCD^18-20^. Raw ECG data may, therefore, be useful to predict SCD since these are routinely collected in any clinical, are low cost and non-invasive^5^. Simple 12-lead ECGs, when incorporated within deep learning frameworks have shown to be able to predict multiple cardiovascular diseases, including cardiomyopathies and heart failure (HF)^21-24^.

The goal of this research was to develop ECG-AI-based models for FCHD risk prediction for the general population that can help clinicians to monitor and assess risk for timely intervention. The specific aims of this research are to i) develop and validate deep learning models that can predict risk for FCHD using both 12-lead and single -lead ECGs ii) assess the concordance of 12-lead vs single lead-ECG only models in FCHD risk prediction and iii) perform time-dependent analysis on FCHD risk to identify time-points when our model is accurate and also allows for timely intervention (e.g. 2-5 year intervals).

## 2. Materials and Methods

### 2.1 Data Sources

Standard 12-lead 10 second time-voltage supine ECG data, demographic, and clinical data and co-morbidities (risk factors) for patients were obtained from the electronic health records (EHR) at the University of Tennessee Health Science Center/Medical Center in Memphis, Tennessee (UTHSC). The study was approved by the IRB of both University of Tennessee Health Science Center, Memphis, TN and Atrium Health Wake Forest Baptist, Winston-Salem, NC.

### 2.2 Outcome Definition

SCD is defined as fatal coronary heart disease (FCHD) which resulted in death due to sudden cardiovascular failure, where the person was otherwise healthy before the event^5,6^. Definite or probable FCHD events were derived from a combination of ICD-9 codes 410, 427.5, 799 and ICD-10 codes I46, I46.2, I46.9, I21, I25.x^5,25^.

### 2.3 Inclusion/Exclusion Criteria

Patients aged 18 or older with at least one ECG recording were included. For initial analysis, we used the ECGs recorded closest to the FCHD event with no restriction on the time between the ECG and FCHD event. For controls, ECGs from the date last seen in the system were used as the anchor point for right censoring.

### 2.4 Electrocardiogram Data

Raw 10-second digital supine 12-lead electrocardiogram data was obtained from the EPIPHANY Cardiology Information System at the University of Tennessee Health Sciences Center, Memphis, TN. The 12-lead ECG data was in either 500Hz or 250Hz voltage. All ECGs were downsampled to 250Hz by removing amplitudes at every other time point. In addition we removed the first second of the 10-second ECG to reduce typical noise associated with ECG initiation. We also developed a single-lead version using ‘lead I’ of the 12-Lead ECG, since this lead is typically mimicked in wearable devices with ECG functionality such as smartwatches. Lead I was used and replicated twelve times, to retain consistency between the model architectures and allowing for streamlined comparison.

### 2.5 Risk Factors

In addition to ECG data, we also included demographic and some clinical characteristics from the EHR that were available to us. Demographic and clinical risk factors included in the study were age, sex, race, diabetes, hypertension, atrial fibrillation (AF), valvular disease (VD), coronary artery disease (CAD), and left ventricular hypertrophy (LVH).

### 2.6 Study Design and Model Development

The data was split in a stratified way into 80% for derivation and 20% as a holdout dataset, which was kept completely independent for final testing and performance reporting. Five-fold cross-validation was employed on the 80% derivation data, resulting in five different models being developed. This data split was kept identical for all the developed models. Each model was assessed using the area under the receiver operating characteristics curve (ROC AUC) for all models and concordance index (C-Index) for Cox Proportional Hazard Regression (CPHR) models. The model performance was assessed based on results on the UTHSC 20% hold-out data. Within the holdout dataset only one ECG (latest ECG) per patient was included.

All model development and analyses were performed using the Python programming language.

### 2.7 Prediction of Fatal Coronary Heart Disease using raw ECGs

This research employed an adjusted ResNet convolutional neural network (CNN) deep learning architecture, outlined in He et al.^26^ and modified by Akbilgic et al. for ECGs^21^, to predict risk of FCHD. The input into this deep learning architecture is a one dimensional (1-D) 12-lead ECG signal and the output was the predicted risk of FCHD. For the single-lead version, the architecture was the same, albeit using different hyperparameters.

### 2.8 Survival analysis with Cox proportional hazard models

Multiple Cox proportional hazard (CPHR) models were developed i) using demographic data, ii) demographics plus clinical data and iii) an additional four models which incorporate the 12- and single-lead ECG-AI output with demographic and clinical (ECG-AI-Cox models; Supplementary Material Table S1). The ECG-AI-Cox models were assessed using the concordance index (C-Index) and time-dependent AUC. Time-dependent AUC is the AUC at each time point from time the ECG was taken until the incident^27^.

### 2.9 Subgroup Analyses

Subgroup analyses were performed for six different age groups, sex, race, and presence of coronary artery disease (CAD), atrial fibrillation (AF) and valvular disease (VD). Statistical significance of the difference between AUCs for between subgroups was assessed using the DeLong’s test.

### 2.10 12- and single-Lead ECG-AI correlation

Correlation between the predictions from the 12- and single-lead ECG-AI models was assessed using coefficient of determination, R^2^, Pearson Correlation, and Spearman Correlation coefficients. The AUCs were statistically compared using the DeLong’s test. In addition, we performed risk stratification on the 12- and single-Lead ECG-AI models to compare risk predictions.

## 3. Results

### 3.1 Clinical Characteristics

The analytical cohort (Table 1) included a total of 167,662 ECGs collected from a total of 50,132 patients after applying inclusion and exclusion criteria. From these, 29,093 were controls with 78,472 ECGs and 21,039 were cases for FCHD with 89,190 ECGs. In some situations, the same visit had multiple ECGs and, therefore, all available ECGs were used. The patient cohort had an average age (age at time the ECG was taken) of 62.58±14.0 (61.63±13.81 for controls and 63.90±14.10 for cases) of which 51.69% were African American (45.50% of controls and 60.24% of cases) and 45.72% were White (51.79% of controls and 37.31% of cases). Within the entire cohort, 53.09% were males (54.04% of controls and 51.78% of cases). The mean time between ECG and FCHD diagnosis was 2.25±2.68 years (median=1.38; minimum=0, maximum=24 years).

**Table 1.** Demographics and clinical characteristics of the patient cohort from UTHSC EHR.

| Demographics/Risk Factors | Total | Controls | Cases |
| --- | --- | --- | --- |
| | $N_{\text{patients}} = 50,132$ | $N_{\text{patients}} = 29,093$ | $N_{\text{patients}} = 21,039$ |
| | $N_{\text{ECGs}} = 167,662$ | $N_{\text{ECGs}} = 78,472$ | $N_{\text{ECGs}} = 89,190$ |
| **Age at ECG taken (years) ±SD** | $62.58 \pm 13.98$ | $61.63 \pm 13.81$ | $63.90 \pm 14.10$ |
| 18-29 | 693 (1.38) | 487 (1.67) | 206 (0.98) |
| 30-39 | 2701 (5.39) | 1,692 (5.82) | 1,009 (4.80) |
| 40-49 | 5809 (11.59) | 3,607 (12.40) | 2,202 (10.47) |
| 50-59 | 11,822 (23.58) | 7,147 (25.57) | 4,675 (22.22) |
| 60-69 | 13,970 (27.87) | 8,293 (28.51) | 5,677 (26.98) |
| $\geq 70$ | 15,769 (31.45) | 8,306 (28.55) | 7,463 (35.47) |
| **Sex** |  |  |  |
| Male (0), n (%) | 26,616 (53.09) | 15,722 (54.04) | 10,894 (51.78) |
| Female (1), n (%) | 23,513 (46.90) | 13,379 (45.96) | 10,143 (48.21) |
| **Race/Ethnicity** |  |  |  |
| African American, n (%) | 25,911 (51.69) | 13,238 (45.50) | 12,673 (60.24) |
| White, n (%) | 22,918 (45.72) | 15,067 (51.79) | 7,851 (37.31) |
| Other or Mixed Race, n (%) | 1303 (2.60) | 788 (2.71) | 515 (2.45) |
| **Diabetes mellitus** |  |  |  |
| No (0), n (%) | 29,842 (59.52) | 20,756 (71.34) | 9,086 (43.19) |
| Yes (1), n (%) | 20,290 (40.47) | 8,337 (28.67) | 11,953 (56.81) |
| **Hypertension** |  |  |  |
| No (0), n (%) | 7,502 (14.96) | 4,238 (14.57) | 3,264 (15.51) |
| Yes (1), n (%) | 42,630 (85.04) | 24,855 (85.43) | 17,775 (84.49) |
| **Left ventricular hypertrophy** |  |  |  |
| No (0), n (%) | 49,889 (99.52) | 29,093 (100) | 20,796 (98.85) |
| Yes (1), n (%) | 243 (0.48) | 0 (0.00) | 243 (1.15) |
| **Valvular disease** |  |  |  |
| No (0), n (%) | 48,834 (97.41) | 28,710 (98.68) | 20,124 (95.65) |
| Yes (1), n (%) | 1,298 (2.59) | 383 (1.32) | 915 (4.35) |
| **Coronary Artery Disease** |  |  |  |
| No (0), n (%) | 7,871 (15.70) | 5,093 (17.51) | 2,778 (13.20) |
| Yes (1), n (%) | 42,261 (84.30) | 24,000 (82.49) | 18,261 (86.80) |
| **Atrial Fibrillation** |  |  |  |
| No (0), n (%) | 37,570 (79.94) | 24,746 (85.06) | 12,824 (60.95) |
| Yes (1), n (%) | 12,562 (25.06) | 4,347 (14.94) | 8,215 (39.05) |

### 3.2 FCHD Risk Prediction Model Evaluations for ECG-AI and ECG-AI-Cox models

The 12- and single-lead ECG-AI models resulted in an AUC of 0.77 (0.76-0.78) and 0.76 (0.76-0.77), respectively on the 20% UTHSC holdout. The D-Cox resulted in a C-Index of 0.60 (0.59-0.61) and AUC of 0.65 (0.63-0.66), while the C-Cox resulted in a C-Index of 0.66 (0.65-0.67) and AUC 0.69 (0.68-0.70). The highest overall AUC of 0.85 (0.84-0.86; bold in Table 2) and C-Index of 0.78 (0.77-0.79) were achieved when 12-Lead ECG-AI predictions were combined with demographic and clinical variables (12-ECG-AI-Cox model), followed by C1-ECG-AI-Cox, achieving an AUC of 0.84 (0.83-0.86) and C-Index of 0.76 (0.75-0.77).

**Table 2.** C-index and/or AUC results of the developed models, evaluated on the 20% holdout set.

| Model Name | UTHSC 20% Holdout |  |
| --- | --- | --- |
|  | C-Index (95% CI) | AUC (95% CI) |
| **12-ECG-AI |  | 0.77 (0.76-0.79) |
| ** 1-ECG-AI |  | 0.76 (0.75-0.77) |
| † D-Cox | 0.60 (0.59-0.61) | 0.65 (0.63-0.66) |
| † C-Cox | 0.66 (0.65-0.67) | 0.69 (0.68-0.70) |
| ^ D12-ECG-AI-Cox | 0.72 (0.71-0.73) | 0.81 (0.80-0.82) |
| ^ D1-ECG-AI-Cox | 0.70 (0.69-0.71) | 0.79 (0.78-0.80) |
| **^ C12-ECG-AI-Cox** | **0.78 (0.77-0.79)** | **0.85 (0.84-0.86)** |
| ^ C1-ECG-AI-Cox | 0.76 (0.75-0.77) | 0.84 (0.83-0.86) |

\*\* ECG-AI Model (Using only ECGs). No C-statistics are reported as these are classification models.
† Cox Proportional hazard (CPHR) models
^ ECG-AI-Cox models (including demographics, clinical and ECG-AI predictions)

The 12-ECG-AI-Cox model (best model) resulted in an accuracy of 78%, sensitivity of 72% and specificity of 82% (Table 3). The coefficients for all CPHR models are provided in Supplementary Material Table S2. It should be noted that within ECG-AI-Cox models, the ECG-AI prediction always had the largest coefficient.

**Table 3.** Confusion matrix for the C12-ECG-AI-Cox model.

| Actual/Predicted | No FCHD | FCHD |  |
| --- | --- | --- | --- |
| No FCHD | 4770 | 1055 | Specificity = 82% |
| FCHD | 1193 | 3000 | Sensitivity = 72% |
|  | Negative Predictive Value = 80% | Positive Predictive Value = 74% | Accuracy = 78% |

### 3.3 Correlation between 12-Lead ECG-AI and single-Lead ECG-AI

When comparing the 12- and 1-ECG-AI models, there was a statistical significance between the two AUCs, resulting in a DeLong p-value < 0.001. However, the prediction results are concordant between the two with a R^2^ of 0.72 (Figure S1), Pearson Correlation Coefficient of 0.85 and Spearman Correlation Coefficient of 0.85.

We also stratified patients at low or high risk for FCHD using both 12- and single-Lead models based on similar specificity and sensitivity (specificity ∽73% and sensitivity of ∽67%) for both models. We compared the stratification results from both models (Table S3). Overall, 84% of the time, both ECG-AI models produced similar class prediction. In further detail, 842 of 5781 people (15%) which were predicted as low risk by 1-ECG-AI were predicted as high risk by the 12-ECG-AI while 717 of 4237 patients (17%) predicted as high risk by 1-ECG-AI were predicted as low risk by 12-ECG-AI.

### 3.4 Time-dependent analysis (ECG-AI-Cox)

Time-dependent analyses of the best model (C12-ECG-AI-Cox) resulted in AUC of 0.91 (0.90-0.92) for 2-year FCHD risk prediction, reducing to AUC=0.84 (0.83-0.85) at 5 years. Accuracy of this model is 84% with a Specificity of 85%, Sensitivity of 83%, PPV of 80% and NPV of 87% (Table 4). A similar trend was obtained for C1-ECG-AI-Cox, but AUC was slightly lower over the first 2 years at 0.89 (0.88-0.90), dropping to AUC= 0.75 (0.74-0.76) after 5 years. We also assessed the ECG-alone models, 12- and 1-ECG-AI, in their power to predict FCHD within 2 years for comparison with the C12-ECG-AI-Cox model. For this, FCHD events within two years were labelled as ‘1’ and any event past two years as ‘0’. Both the 12- and 1-ECG-AI models slightly increased in AUC from 0.77 to 0.80 (0.79-0.81) and AUC of 0.76 to 0.78 (0.77-0.80), respectively.

**Table 4.** Confusion matrix for the 2-year C12-ECG-AI-Cox prediction model.

| Actual/Predicted | No FCHD | FCHD |  |
| --- | --- | --- | --- |
| No FCHD | 4936 | 889 | Specificity = 85% |
| FCHD | 715 | 3478 | Sensitivity = 83% |
|  | Negative Predictive Value = 87% | Positive Predictive Value = 80% | Accuracy = 84% |

### 3.4 Subgroup Analyses

We performed subgroup analysis on age groups, sex and race/ethnicity and diagnosis for CAD using the best performing ECG-AI-Cox (Table 5). Subgroup analysis showed no significant difference when comparing males and females with AUCs of 0.79 (0.77-0.81) and 0.80 (0.78-0.82) and p=0.356. The same AUCs were achieved when comparing African American vs white races, with a p-value=0.393. There was also no significant difference in AUCs for subgroups with and without CAD with p=0.397, with and without VD with p =0.187) and with and without AF (p=0.187).

**Table 5.** Subgroup analysis on sex, age groups, race, ethnicity and CAD using the best operating model (C12-ECG-AI-Cox). Comparisons were performed using the DeLong test. *Detailed subgroup analysis comparing all age groups are provided in supplementary material Table S4.

| Subgroups | ECG-AI-Cox | DeLong Test p-value |
| --- | --- | --- |
| *Sex* |  |  |
| Male | 0.80 (0.79-0.81) | 0.4265 |
| Female | 0.79 (0.78-0.81) |  |
| *Age\** |  |  |
| 18-29 | 0.82 (0.73-0.91) | Only significant difference is between 40-49, 50-59 and 60-69 vs >70 (see Table S4) |
| 30-39 | 0.83 (0.78-0.87) |  |
| 40-49 | 0.83 (0.80-0.85) |  |
| 50-59 | 0.82 (0.80-0.83) |  |
| 60-69 | 0.81 (0.79-0.83) |  |
| ≥70 | 0.77 (0.75-0.79) |  |
| *Race* |  |  |
| African American | 0.80 (0.78-0.81) | 0.371 |
| White | 0.79 (0.77-0.80) |  |
| *Coronary Artery Disease (CAD)* |  |  |
| Yes | 0.80 (0.79-0.81) | 0.397 |
| No | 0.79 (0.77-0.81) |  |
| *Atrial Fibrillation (AF)* |  |  |
| Yes | 0.76 (0.74-0.78) | 0.187 |
| No | 0.78 (0.76-0.79) |  |
| *Valvular Disease (VD)* |  |  |
| Yes | 0.75 (0.69-0.82) | 0.187 |
| No | 0.80 (0.79-0.80) |  |

Subgroup results (Tables 5 and Table S4) for different age groups showed that the highest AUC of 0.83 was achieved for age groups 30-39 and 40-49 with no significant difference between these groups. The only significant differences were found when comparing AUCs of the 40-49 vs **≥**70 (AUC = 0.77), 50-59 (AUC = 0.82) vs **≥**70 and 60-69 (AUC = 0.81) vs **≥**70 age groups with p-values of <0.001.

## 4. Discussion

SCD is sudden cessation of cardiac activity with hemodynamic collapse^28^ and predominantly occur in patients with structural heart disease, especially coronary heart disease and fatal ventricular arrhythmias. As a result, FCHD is often used as a proxy to determine SCD, since the latter’s true identification is difficult to ascertain^5,6^. The most important problem with SCD is the suddenness and rapidity with which the event is fatal. Therefore, there is a need for prediction models that can be easily accessible and used within routine clinical care to increase monitoring and follow-ups. In this research, we developed and compared multiple models to assess the power of ECGs to predict risk of FCHD over time. Results show that the highest AUC was obtained using C12-ECG-AI-Cox (AUC = 0.85). Time-dependent analysis resulted in an AUC of 0.91 in predicting 2-year risk for FCHD. Furthermore, the 12-ECG-AI and 1-ECG-AI have moderate, yet similar, AUCs with high correlation between the predictions. This enforces the notion that ECG-based predictions are very important in determining FCHD risk, increasing the potential of usability for monitoring and smart wearables.

While some time-dependent FCHD and SCD analyses have been previously performed, high accuracy was achieved up to 24hrs before and moderate to low accuracy up to 10 years before^5^. Kwon et al.^20^ demonstrated that in-hospital sudden cardiac arrest could be predicted in >50% of patients several hours before the event using machine learning. However predicting SCD in such a short time period may not be clinically helpful for intervention. Therefore, using simple and routinely-collected data, e.g. ECG, can be an asset within AI models to increase pre-screening efforts of fatal events. In a large-scale prospective study, it was demonstrated that deep learning artificial intelligence could successfully predict cardiac arrest using diverse formats of ECGs, mostly depending on ECG-based features and data of amplitude and duration of individual ECG waves^20^. However, the use of numerous ECG features can be difficult to obtain, and might not always be available from clinical institutes, increasing the need for ECG-AI methods using raw ECGs in predicting outcomes.

In some published AI-driven FCHD or SCD prediction research, very high accuracies (AUC>0.85) were achieved hours before the event, which reduces usability for therapeutic interventions as well as they are not applicable to population level screening^5,15,29^. However, results from our research show sustained high AUC of 0.91 for 2-year risk prediction of FCHD. The C1-ECG-AI-Cox resulted in very similar time-dependent AUC of 0.89 within the same timeline. In addition, we also show that ECG-based predictions substantially improve model predictions when compared to models developed using only demographics or demographics plus clinical data. This highlights the importance of ECG-based models and assessments years before the potential SCD incident. In this period, the model can clearly distinguish between FCHD and non-FCHD events (NPV = 87%, PPV = 80%), making it increasingly useful if incorporated within clinical settings as a tool for risk assessment.

Furthermore, following the work in this research on feasibility of a single-lead ECG-AI model, results show that 12- and single-Lead ECG-based predictions are very correlated (Figure S1), and while the AUC is slightly lower in the single-lead based models, it paves the way for the integration within mobile AI platforms such as ECG-Air^30^ that can gather ECG from wearable devices such as smart watches for easier, simpler and cost-effective pre-screening at the population level. The availability of tools for pre-screening is important since SCD is highly correlated with certain cardiac diseases such as HCM, the larger subset of people with LVH and some risk factors (e.g. diabetes) ^31-33^, which all showed high coefficients in the CPHR models developed in this research. Having a simple and usable AI model that can be integrated within a smart device, especially a smart watch with single-Lead ECG capabilities, can be an asset to the clinical workflow and help decision making for triaging, testing and risk assessment.

### Limitations and future studies

This study had some limitations. Predominantly, while FCHD has been reported to be a proxy of SCD, true SCD event data could possibly improve the accuracy of the models and therefore, future work will focus on testing and validating our models on SCD outcomes. In addition, our current efforts are focusing on gathering data a larger set of data from additional EHRs and established NHLBI-funded cohort studies for both training and external validation. Although we assessed the correlation of 12-lead vs single-lead models, as a feasibility study for future remote monitoring via wearables, we did not use ECGs obtained from wearable ECG devices (e.g. smartwatches). Our preliminary work shows that predictions from single-lead clinical ECGs and ECGs from smartwatches are also highly correlated and we will expand on this research for FCHD and SCD. Additional future studies lie in designing an AI-assisted randomized clinical trial to assess the effectiveness of AI-assisted follow-up protocols on patients predicted at high SCD and compare outcomes to standard of care.

## Conclusion

The results from this research indicate that FCHD/SCD events can be predicted from both single- and 12-lead ECGs using state-of-the-art methods. The deep learning models applied to ECG data can be combined with clinical data using a survival analysis framework to analyze time-dependent risks and allow for more patient-specific data commonly available in electronic health records. This research shows that both 12- and single-Lead ECG-based models operate similarly and, therefore, this not only serves as a proof-of-concept for FCHD and SCD prediction but as a large contribution for the integration within e-health and remote monitoring systems, which can help directly target people within the general population. High 2-year prediction accuracy can guide developing novel protocols for detailed examination and close monitoring of those identified at high risk.

## Data Availability

Data used was obtained from the University of Tennessee Health Science Center under a Data User Agreement. Final AI models will be shared on GitHub after manuscript publication.

## References

1. Tsao CW, Aday AW, Almarzooq ZI, Anderson CA, Arora P, Avery CL, et al. Heart disease and stroke statistics—2023 update: a report from the American Heart Association. Circulation. 2023;147(8):e93–e621.

2. Benjamin EJ, Virani SS, Callaway CW, Chamberlain AM, Chang AR, Cheng S, et al. Heart disease and stroke statistics—2018 update: a report from the American Heart Association. Circulation. 2018;137(12):e67–e492.

3. Srinivasan NT, Schilling RJ. Sudden cardiac death and arrhythmias. Arrhythmia & electrophysiology review. 2018;7(2):111.

4. Chugh SS, Reinier K, Teodorescu C, Evanado A, Kehr E, Al Samara M, et al. Epidemiology of sudden cardiac death: clinical and research implications. Progress in cardiovascular diseases. 2008;51(3):213–28.

5. Perez-Alday EA, Bender A, German D, Mukundan SV, Hamilton C, Thomas JA, et al. Dynamic predictive accuracy of electrocardiographic biomarkers of sudden cardiac death within a survival framework: the Atherosclerosis Risk in Communities (ARIC) study. BMC cardiovascular disorders. 2019;19(1):1–19.

6. Bogle BM, Sotoodehnia N, Kucharska-Newton AM, Rosamond WD. Vital exhaustion and sudden cardiac death in the Atherosclerosis Risk in Communities Study. Heart. 2018;104(5):423–9.

7. Wallace E, Howard L, Liu M, O’Brien T, Ward D, Shen S, et al. Long QT syndrome: genetics and future perspective. Pediatric cardiology. 2019;40:1419–30.

8. Angelini P. Coronary artery anomalies: an entity in search of an identity. Circulation. 2007;115(10):1296–305.

9. Gentile F, Castiglione V, De Caterina R. Coronary artery anomalies. Circulation. 2021;144(12):983–96.

10. John RM, Tedrow UB, Koplan BA, Albert CM, Epstein LM, Sweeney MO, et al. Ventricular arrhythmias and sudden cardiac death. The Lancet. 2012;380(9852):1520–9.

11. Marijon E, Garcia R, Narayanan K, Karam N, Jouven X. Fighting against sudden cardiac death: need for a paradigm shift-Adding near-term prevention and pre-emptive action to long-term prevention. Eur Heart J. 2022;43(15):1457–64.

12. Shameer K, Johnson KW, Glicksberg BS, Dudley JT, Sengupta PP. Machine learning in cardiovascular medicine: are we there yet? Heart. 2018;104(14):1156–64.

13. Soudan B, Dandachi FF, Nassif AB. Attempting cardiac arrest prediction using artificial intelligence on vital signs from Electronic Health Records. Smart Health. 2022:100294.

14. O’Mahony C, Jichi F, Pavlou M, Monserrat L, Anastasakis A, Rapezzi C, et al. A novel clinical risk prediction model for sudden cardiac death in hypertrophic cardiomyopathy (HCM risk-SCD). European heart journal. 2014;35(30):2010–20.

15. Shiraishi Y, Goto S, Niimi N, Katsumata Y, Goda A, Takei M, et al. Improved prediction of sudden cardiac death in patients with heart failure through digital processing of electrocardiography. EP Europace. 2023.

16. Konety SH, Koene RJ, Norby FL, Wilsdon T, Alonso A, Siscovick D, et al. Echocardiographic predictors of sudden cardiac death: the atherosclerosis risk in communities study and cardiovascular health study. Circulation: Cardiovascular Imaging. 2016;9(8):e004431.

17. De Lio F, Andreis A, De Lio G, Bellettini M, Pidello S, Raineri C, et al. Cardiac imaging for the prediction of sudden cardiac arrest in patients with heart failure. Heliyon. 2023.

18. Chen Y-Y, Chung F-P, Lin Y-J, Chien K-L, Chang W-T. Exploring the Risk Factors of Sudden Cardiac Death Using an Electrocardiography and Medical Ultrasonography for the General Population Without a History of Coronary Artery Disease or Left Ventricular Ejection Fraction< 35% and Aged> 35 Years-A Novel Point-Based Prediction Model Based on the Chin-Shan Community Cardiovascular Cohort-. Circulation Journal. 2022;87(1):139–49.

19. Brugada J, Brugada R, Brugada P. Determinants of sudden cardiac death in individuals with the electrocardiographic pattern of Brugada syndrome and no previous cardiac arrest. Circulation. 2003;108(25):3092–6.

20. Kwon J-m, Kim K-H, Jeon K-H, Lee SY, Park J, Oh B-H. Artificial intelligence algorithm for predicting cardiac arrest using electrocardiography. Scandinavian journal of trauma, resuscitation and emergency medicine. 2020;28(1):1–10.

21. Akbilgic O, Butler L, Karabayir I, Chang PP, Kitzman DW, Alonso A, et al. ECG-AI: electrocardiographic artificial intelligence model for prediction of heart failure. European Heart Journal-Digital Health. 2021;2(4):626–34.

22. Verbrugge FH, Reddy YN, Attia ZI, Friedman PA, Noseworthy PA, Lopez-Jimenez F, et al. Artificial Intelligence Predicts Atrial Fibrillation Development from the 12-lead Electrocardiogram in Heart Failure with Preserved Ejection Fraction. Journal of Cardiac Failure. 2020;26(10):S76.

23. Gunturkun F, Davis RL, Armstrong GT, Jefferies JL, Ness KK, Green DM, et al. Deep learning for improved prediction of late-onset cardiomyopathy among childhood cancer survivors: A report from the St. Jude Lifetime Cohort (SJLIFE). Journal of Clinical Oncology. 2020;38(15_suppl):10545-.

24. Guerrier K, Gunturkun F, Wetzel G, Akbilgic O, Davis R, Towbin J. Human versus machine: does artificial intelligence add value to identification of hypertrophic cardiomyopathy in pediatric patients? European Heart Journal. 2022;43(Supplement_2).

25. White AD, Folsom AR, Chambless LE, Sharret AR, Yang K, Conwill D, et al. Community surveillance of coronary heart disease in the Atherosclerosis Risk in Communities (ARIC) Study: methods and initial two years’ experience. Journal of clinical epidemiology. 1996;49(2):223–33.

26. He K, Zhang X, Ren S, Sun J, editors. Deep residual learning for image recognition. Proceedings of the IEEE conference on computer vision and pattern recognition; 2016.

27. Kamarudin AN, Cox T, Kolamunnage-Dona R. Time-dependent ROC curve analysis in medical research: current methods and applications. BMC medical research methodology. 2017;17(1):1–19.

28. Kannel WB, Schatzkin A. Sudden death: lessons from subsets in population studies. Journal of the American College of Cardiology. 1985;5(6):141B–9B.

29. Mirhoseini SR, JahedMotlagh MR, Pooyan M, editors. Improve accuracy of early detection sudden cardiac deaths (SCD) using decision forest and SVM. Proceedings of the International Conference on Robotics and Artificial Intelligence; 2016.

30. McCraw CA, Karabayir I, Akbilgic O. ECG-AIR: AN AI PLATFORM FOR REMOTE SMARTWATCH ECG-BASED CARDIOVASCULAR DISEASE DETECTION AND PREDICTION. Cardiovascular Digital Health Journal. 2022;3(4):S7.

31. Schirmer H, Lunde P, Rasmussen K. Prevalence of left ventricular hypertrophy in a general population; The Tromsø Study. European heart journal. 1999;20(6):429–38.

32. Norrish G, Ding T, Field E, Cervi E, Ziólkowska L, Olivotto I, et al. Relationship between maximal left ventricular wall thickness and sudden cardiac death in childhood onset hypertrophic cardiomyopathy. Circulation: Arrhythmia and Electrophysiology. 2022;15(5):e010075.

33. Lynge TH, Svane J, Pedersen-Bjergaard U, Gislason G, Torp-Pedersen C, Banner J, et al. Sudden cardiac death among persons with diabetes aged 1–49 years: a 10-year nationwide study of 14 294 deaths in Denmark. European heart journal. 2020;41(28):2699–706.

